# Low dose hydroxychloroquine prophylaxis for COVID-19 – a prospective study

**DOI:** 10.1101/2021.09.13.21262971

**Authors:** RE-HCP2 COVID study group, Mayank Agarwal, Rajat Ranka, Prasan Kumar Panda, Ajay Kumar, Gaurav Chikara, Suresh K Sharma, Rizu Negi, Ramanuj Samanta, Rohit Walia, Yogesh Arvind Bahurupi, Sarama Saha, Minakshi Dhar, Prakhar Sharma, Arvind Kumar Gupta, UB Mishra, Manoj Kumar Gupta, Ravi Kant

## Abstract

**Background:** Since the outbreak of COVID-19 pandemic, the world began a frantic search for possible prophylactic options. While trials on hydroxychloroquine (HCQ) prophylaxis are ongoing, concrete evidence is lacking. The study aimed to determine the relative efficacy of various doses of oral HCQ in prophylaxis and mitigating the severity of COVID-19 in healthcare workers.

**Methods:** This was a prospective cohort with four arms (high, medium, low dose, and control) of HCQ prophylaxis, used by healthcare workers at a tertiary care center in India. Participants were grouped as per their opting for any one arm on a voluntary basis as per institute policy under the Government guidance. The outcomes studied were COVID-19 positivity by RT-PCR and its severity assessed by WHO COVID-19 severity scale.

**Results:** Total 486 participants were enrolled, of which 29 (6%) opted for low dose, 2 (<1%) medium dose, and none for high dose HCQ while 455 (93.6%) were in the control arm. Of the 164 participants who underwent RT-PCR, 96 (58.2%) tested positive. Out of these 96 positive cases, the majority of them (79 of 96 [82.3%]) were ambulatory and were managed conservatively at home. Only 17.7% (17 of 96) participants, all of them from the control group, required hospitalization with the mild-moderate disease. None of the participants had severe disease, COVID-related complications, ICU stay, or death. The difference in the outcome assessed amongst the various arms was statistically insignificant (p value >0.05).

**Conclusion:** This single-center study demonstrated that HCQ prophylaxis in healthcare workers does not cause a significant reduction in COVID-19 as well as mitigating its severity in those infected. At present, most of the trials have not shown any benefit. The debate continues to rage, should HCQ prophylaxis be given to healthcare workers for chemoprophylaxis?

## Introduction

Since late 2019, a health crisis created by the novel severe acute respiratory syndrome coronavirus-2 (SARS-CoV-2) has gripped the entire world, taking a massive toll on people and nations. It has inflicted substantial societal and economic devastation and has overwhelmed healthcare systems. Directly or indirectly, the virus has had a toll on almost the entire population of the world, at a scale not seen in over a century. It has also left considerable strain on the healthcare system, with frontline healthcare workers at a high risk of contracting the infection.

Since the onset of the covid outbreak, the world began a frantic search for possible therapeutic options. Given that hydroxychloroquine has been in use for decades with a good safety profile, there has been a lot of focus regarding its potential use against COVID-19. Many large-scale trials have been launched evaluating the efficacy of hydroxychloroquine as a therapeutic as well as a prophylactic option. An early study from China reported the in-vitro efficacy of chloroquine against the novel coronavirus.(1) It is thought to impair the terminal glycosylation of the angiotensin-converting–enzyme 2 (ACE2) receptor, which is the binding site for the envelope spike glycoprotein and has been shown to inhibit endolysosome function.(2) The pharmacokinetics of hydroxychloroquine, such as its high lung concentration (500 times the blood concentration) and the long half-life are ideally suited for its use as an agent for prophylaxis.(3)

Subsequently over the next few months, the results of trials ruled out any benefit with the use of hydroxychloroquine (HCQ) for the treatment of COVID 19,(4)(5) and trials involving HCQ as a treatment option were discontinued. However, the results of the trials on prophylaxis have been ambiguous with few of them showing conflicting outcomes. Nonetheless, few have postulated that the postexposure and early treatment trials may not have achieved therapeutic concentrations early enough to demonstrate any benefit.(6,7) The Indian Council of Medical Research has recommended chemoprophylaxis with HCQ (400 mg twice on day 1, then 400 mg once a week thereafter for 7 weeks) nationally for asymptomatic healthcare workers at high risk for COVID-19, despite no substantial evidence favouring its use as chemoprophylaxis.(8) Even a year after the pandemic began, the question regarding the efficacy of HCQ as possible prophylaxis remains unanswered. Hopefully, in the next few months, as more and more trials like ours come into the public domain, that answer may be clearer.

## Material and methods

### Study design

We conducted a pragmatic prospective multiple arms cohort study to determine the relative efficacy of various hydroxychloroquine (HCQ) doses in prevention and mitigation of severity of symptomatic COVID-19 disease in high-risk healthcare workers at a tertiary care hospital in India. Participants were educated regarding the various dose schedules of HCQ. They were asked to take an informed decision regarding enrolment into one of the four study arms after a valid prescription. There was no randomization since the decision for drug prophylaxis and dosing was taken by the participants themselves as per available Government guideline and other literatures.

This study was approved by the Institutional Ethics Committee (IEC) at All India Institute of Medical Sciences, Rishikesh (CTRI/2020/06/025593). The study enrolment began on April 12, 2020, and ended on June 7, 2020, while follow-up was completed by October 26, 2020.

### Participants

We included asymptomatic healthcare workers (HCWs) with high risk of exposure to COVID-19 infection. High risk of exposure was defined as all HCW’s including residents, nurses, paramedics, and attending staff who had direct contact with COVID-19 patients in the emergency department, COVID dedicated wards, operating rooms, intensive care units (ICUs), or those who documented having contact with COVID-19 patients during their work. HCWs who were reluctant to take any prophylaxis; or those with history of any of the following conditions: retinopathy or retinal disease, cardiomyopathy, cardiac arrhythmia, prolonged QT syndrome, psoriasis, porphyria cutanea tarda, epilepsy, myasthenia gravis, myopathy of any cause, serious hepatic or renal disease, glucose-6-phosphate dehydrogenase deficiency, severe depression; or those taking medication with known serious hepatotoxic effects or known interaction with HCQ were included in the control arm of the study.

We excluded participants with weight outside range (45 kg-150 kg); prior enrolment into this study; active or previous COVID-19 diagnosis within the last 6 months; or self-reported current acute respiratory infection; or inability/unwillingness to be followed up for the trial period.

### Study setting

All eligible HCWs employed at the hospital were enrolled in the study. Consent was obtained electronically after the participants read the online information sheet regarding the nature and implications of the study. The participants were free to withdraw from the study at any time for any reason without prejudice to future care and without any obligation to give the reason for withdrawal. However, they were educated for the importance of daily monitoring and to continue participating in the study.

At the time of enrolment, we collected demographic information, comorbidities, and relevant medical history of all participants along with general and physical examination. Blood investigations including complete blood count, renal and liver function tests, and G6PD levels were measured. A baseline electrocardiogram and ophthalmological evaluation were also done.

### Intervention

Participants were educated regarding the different prophylaxis dose schedules of HCQ and asked to take a voluntary decision for enrolment into one of the four arms of the study: ICMR/low-dose (400mg HCQ twice daily on Day 1 followed by 400mg weekly for 7 weeks) or Medium-dose (400mg HCQ once daily for 4 days followed by twice weekly for 3 months) or High-dose (400mg HCQ once daily for 4 days followed by 200 mg daily for 3 months) or Control arm which included HCWs with any contraindication to HCQ or they did not opt for any prophylaxis.

### Study Assessments

Follow up and assessment of participants were done through a combination of telephonic interviews as well as self-reporting on a web-based software (@Zifo RnD Solutions, Chennai, India). Participants used a secure clinical registry form (eCRF) login to self-record a daily census with questions regarding symptoms, exposure, and treatment adherence. They were sent a reminder through text message by the software on their mobile phones with a link to the website. Along with self-reporting all participants were required to physically report symptoms, general wellbeing, and compliance at enrolment followed by on day 30, 60, 90 or whenever they completed prophylaxis. In case of participant reporting COVID-19 related symptoms, they were asked to undergo RT-PCR testing, if tested positive was managed according to the institute treatment protocol.

The study participants were monitored for two months post completion of prophylaxis or development of confirmed symptomatic COVID-19 infection to determine the outcome in terms of recovery/severity/mortality. All adverse events were recorded with clinical symptoms and accompanied with a simple, brief description of the event. Each adverse event was assessed for severity, causality, seriousness, and expectedness.

### Outcomes

The primary outcomes were measured in terms of incidence of COVID-19 in each one of the four arms and the peak severity of COVID-19 in the positive study participants over the study period. Confirmed COVID-19 was defined as participant with reverse transcriptase polymerase chain reaction (RT-PCR) positive for SARS-CoV-2. Severity was graded on the Ordinal WHO COVID-19 severity scale. These outcome definitions were based on WHO R&D Blueprint consensus definitions for COVID-19.(9)

Secondary outcomes included incidence of COVID-19 related complications such as respiratory failure requiring intubation, acute respiratory distress syndrome, delirium, shock requiring inotropes, sepsis, acute kidney injury and acute liver injury, duration of ICU stay, and mortality.

### Statistical analysis

Summary of the baseline characteristics of all participants was estimated by frequency and percentage for categorical variables whereas mean and standard deviation (or median and inter-quartile range for non-normally distributed data) for continuous variables. The primary endpoint of incidence of COVID19 was presented as proportion of participants reported with RTPCR positive test. Association between RT-PCR positivity and COVID-19 severity was estimated by Chi-Square test. Comparison between the means of COVID-19 severity scores among the two arms was done using Mann Whitney test.

## Results

### Basic characteristics

We enrolled 486 adult asymptomatic HCWs out of which 29 (6%) participants opted for low-dose HCQ, 2 (<1%) participants for medium-dose HCQ and none for high-dose HCQ while 455 (93.6%) participants were enrolled in the control group as they did not opt for any prophylaxis or had contraindication to HCQ use. Since only 2 participants opted for medium dose prophylaxis they were excluded from the results for statistical reasons (Figure 1). The demographic and clinical characteristics of the participants were comparable among arms (Table 1). No adverse events were reported in either of the study arms.

**Figure 1:**
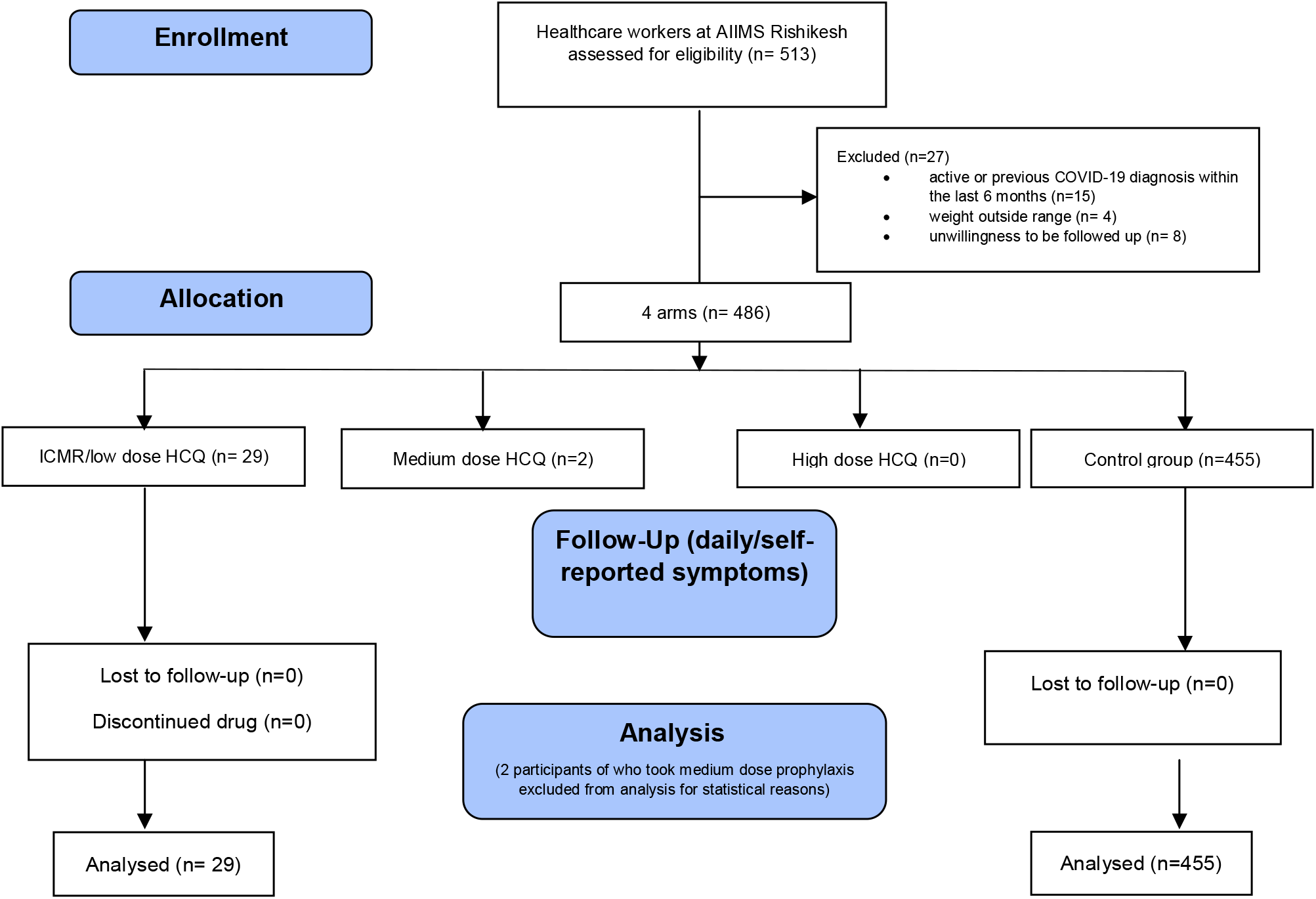
The study flow

**Table 1:** Distribution of Study participants according to demographic and clinical profile

| Variables | Low dose hydroxychloroquine<br>(N=29)<br>(Mean±SD/ n, %) | Control<br>(N=455)<br>(Mean±SD/ n, %) | P value |
| --- | --- | --- | --- |
| **Gender** |  |  | 0.976 |
| Male | 20 (69.0) | 315 (69.2) |  |
| Female | 9 (31.0) | 140 (30.8) |  |
| **Age (years)** | 30.2±6.6 | 27.6±4.2 | 0.02 |
| **Occupation** |  |  |  |
| Attending staff | 0 (0.0) | 37 (8.1) |  |
| Nurse | 13 (44.8) | 320 (70.3) |  |
| Paramedic | 2 (6.9) | 20 (4.4) |  |
| Resident doctors | 14 (48.3) | 78 (17.1) |  |
| **Height (cms)** | 166.6±10.5 | 167.1±8.6 | 0.788 |
| **Weight (kgs)** | 71.7±12.5 | 68.0±12.2 | 0.123 |
| **BMI (kg/m<sup>2</sup>)** | 25.7±2.8 | 24.3±3.5 | 0.041 |
| **H/o allergies** |  |  | 0.126 |
| Yes | 4 (13.8) | 29 (6.4) |  |
| No | 25 (86.2) | 426 (93.6) |  |
| **Co-morbidities** |  |  |  |
| Hypertension | 5 (17.2) | 12 (2.6) | 0.002 |
| Diabetes mellitus | 0 (0) | 4 (0.9) | 1.00 |
| Chronic kidney disease | 0 (0) | 1 (0.2) | 1.00 |
| Coronary artery disease | 0 (0) | 3 (0.6) | 1.00 |
| COPD | 0 (0) | 2 (0.4) | 1.00 |
| Obstructive sleep apnoea | 0 (0) | 3 (0.7) | 1.00 |
| Asthma | 0 (0) | 11 (2.4) | 1.00 |
| Retinal pathology | 0 (0) | 2 (0.4) | 1.00 |
| **Concomitant medications** | 5 (17.2) | 40 (8.7) | 0.067 |

**Table 2:** Outcomes of hydroxychloroquine therapy

**Figure 2: Distribution of study participants according to their COVID-19 status**
| Table 2: Outcomes of hydroxychloroquine therapy |  |  |  |  |
| --- | --- | --- | --- | --- |
| Outcome |  | Low dose hydroxychloroquine (N=14) (Mean±SD/ n, %) | Control (N=150) (Mean±SD/ n, %) | P value |
| COVID status (RT-PCR) |  |  |  |  |
| • Positive |  | 6 (42.9) | 90 (60) | 0.213 |
| • Negative |  | 8 (57.1) | 60 (40) |  |
| COVID-19 Severity Scale |  | 1.17±0.5 | 1.6±0.8 | 0.213 |
| Ambulatory |  | 6 (100) | 73 (81.1) | 0.587 |
| Hospitalised mild-mod disease |  | 0 | 17 (18.9) |  |

### Outcomes

The overall incidence of confirmed COVID-19 was 20.7% (6 of 29) in the low-dose arm and 19.8% (90 of 455) in the control arm (Figure 2). The association between COVID-19 incidence and dosing in the two arms was found to be statistically insignificant. The mean of the severity of COVID-19 in the positive study participants was lesser in the low dose arm (1.17±0.4.8) as compared to the control arm (1.6±0.818), however the difference in means of severity scores in the two dosing arms was statistically insignificant. Out of these 96 positive cases, majority of them were ambulatory and were managed conservatively at home. Only 17.7% (17 of 96) participants, all of them from control group, required hospitalization with mild-moderate disease.

**Figure 2:**
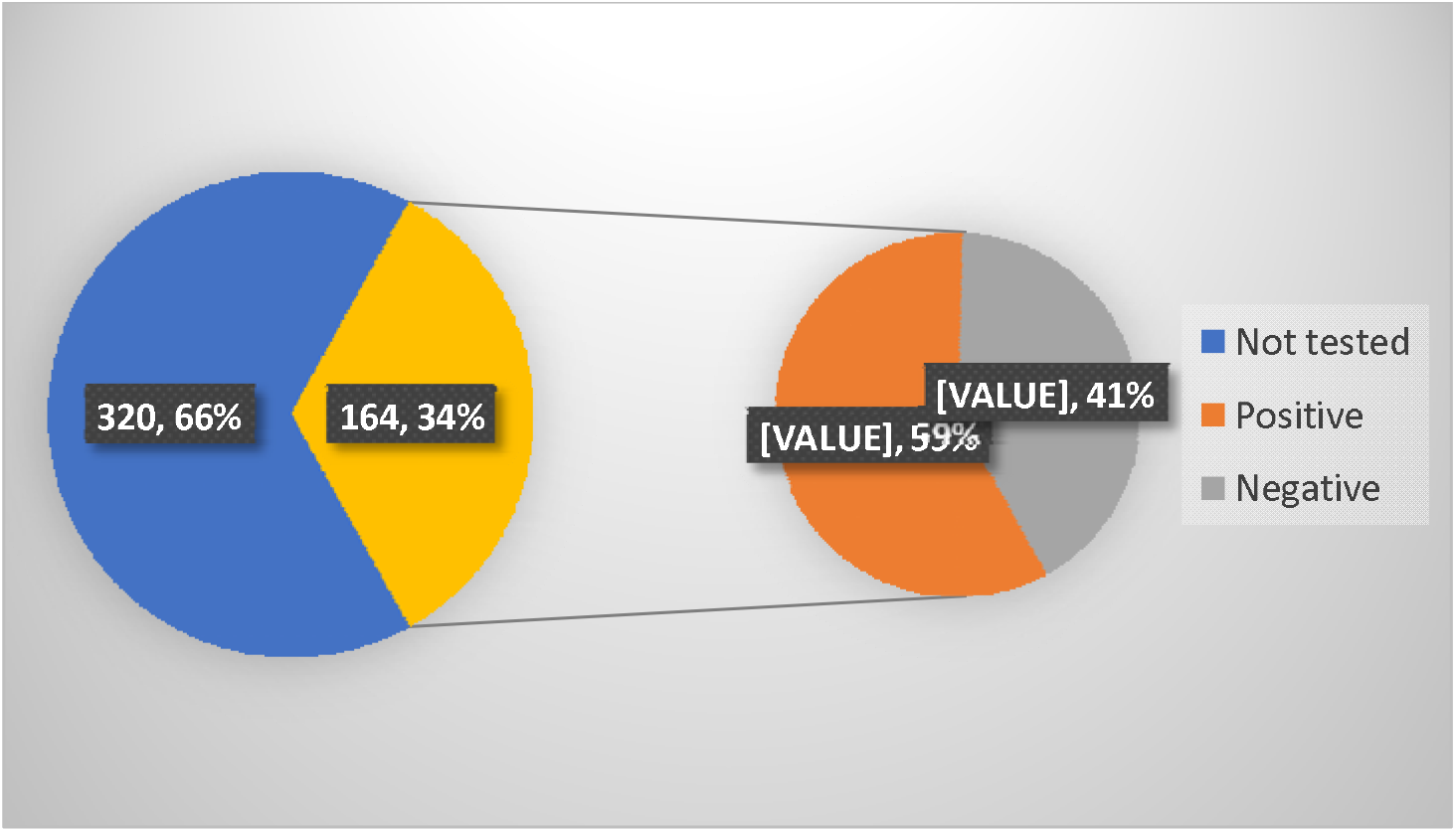
Distribution of study participants according to their COVID-19 status.

## Discussion

In this cohort study, evaluating hydroxychloroquine (HCQ) for the pre-exposure prophylaxis of COVID-19 in healthcare workers (HCWs), we found no statistically significant reduction in the incidence of COVID-19 in those receiving the 400 mg weekly dose of HCQ compared to controls. We also intended to evaluate the prophylactic efficacy of medium and high-dose hydroxychloroquine, but they were excluded since only two participants were enrolled.

Reason for no benefit with HCQ may be due to it being ineffective in vivo or the drug concentration being too low.(10) Although we did not measure the serum concentrations of HCQ, but in a similar study done by Rajasingham et al., they found no difference in the drug concentrations between those who developed COVID-19 and those who did not.(11) The ICMR dose (400 mg once weekly HCQ) is predicted to achieve plasma concentration that is more than the in-vitro half maximal effective concentration (EC_50_).(12) However many studies have found that there is a discrepancy in the simulated and observed drug concentrations and the reason may lie in the sequestration of drug in the leukocytes and platelets.(11,13) Taking this correction into consideration, the extrapolated plasma concentrations are unlikely to reach EC_50_ even with daily dosing of HCQ.(11) This underscores the need for drug concentration measurement in any future or ongoing trial evaluating the efficacy of HCQ in the context of COVID-19. The present study shows that prophylaxis with 400 mg of HCQ weekly is ineffective, but it is doubtful whether even a significantly higher dose would have been any better. In one double blind randomised trial of 125 patients, even 600 mg of daily HCQ did not reduce the incidence of COVID-19 in HCWs.(14) Even in a pre-clinical investigation in COVID-19 macaque model, there was no difference in the antiviral activity with varied doses of hydroxychloroquine.(10)

Apart from the above-mentioned pre-exposure trials of HCQ, there have been a few other randomized controlled trials that have evaluated HCQ for post-exposure prophylaxis. Most of these studies did not report any evidence of efficacy and showed higher adverse effects with the use of HCQ.(7,15,16) The PEP-CQ study did show a decreased incidence of COVID-19 in the HCQ group, but the design was open-label and the sample size was small.(17)

When compared to the controls, those who opted for HCQ had a higher age (30.2 vs 27.6 years, p-value 0.02), prevalence of hypertension (17.2 % vs 2.6 %, p-value 0.002) and body mass index (25.67 vs 24.3, p-value 0.04). Despite these risk factors, those who took the drug developed less severe disease (ordinal scale score 1.17 vs 1.6), but the difference failed to reach statistical significance. Infact all the 17 participants who required hospitalization for COVID-19 had not taken HCQ prophylaxis. Apart from the initial months, enrolling participants further was difficult, especially in the intervention arms. Later onwards, the enrolment declined precipitously. This was mainly driven by safety concerns flagged by a few reports, FDA warning, and emerging literature regarding the lack of treatment efficacy of HCQ in trials worldwide.(5,18,19) No adverse effects were reported by any of the 29 participants who took 400 mg weekly HCQ, and the drug was well tolerated. None of the participants on follow up showed any abnormity in electrocardiogram and fundus examination This study had few limitations. Firstly, the design of the study was a cohort study, just observing the participants taking HCQs or not. The sample size in the intervention arm was very small and we cannot exclude the possibility of a modest prophylactic effect that remained undetected. Moreover, the frequency of exposure to COVID-19 was not quantified and the drug concentrations were not monitored. The cohort comprised mostly of young and healthy healthcare workers. Therefore, the results cannot be generalised for the more vulnerable sections of the population. The study was conducted at a single centre in the North India and therefore, may not be representative of disease prevalence and exposure in other regions. The design of the study was such that the participants got themselves tested as per the clinical need only and as a result, 66 % of the total participants were never tested during the study period. Hence, asymptomatic COVID-19 infections were not accounted for.

Drugs like ivermectin and antiretroviral agents have also been tried and tested for the prevention of COVID-19, but head-to-head trials have been lacking. Of these, ivermectin received governmental approval for mass distribution in various states in India.(20) However, as per the available evidence, none of these agents should be used for chemo-prophylaxis outside the context of a clinical trial.(21,22)

Ever since the mass vaccinations roll out, an effective means of chemo-prophylaxis against the SARS-CoV2 may no longer be a critical need in the developed countries. However, in many low- and middle-income countries including India, the vaccination coverage is far from satisfactory and therefore, the search for a safe and reasonable chemoprophylaxis should continue until a large population of individuals gets vaccinated.

## Data Availability

After obtaining approval from corresponding author, de-identified data can be shared.

## Authors’ contributions

MA, RR, GC, RN, RS, RW, SS and YAB contributed to the data collection, analysis, and was involved in manuscript writing. PKP, AK, SKS, MD, PS, AKG, UBM, MKG, and RK gave the concept, critically reviewed the draft, approved for publication along with all authors including agreed to be accountable for all aspects of the work.

## Ethics and Data sharing

The study was done after institute ethical approval and as per declaration of Helsinki. After obtaining approval from corresponding author, de-identified data can be shared.

## Acknowledgment

A gratitude to the entire COVID-19 management team and participants during this pandemic crisis for helping in data collection. A special thanks to Zifo RnD Solutions, Chennai, India for providing data software used for data management. Also thanks to Dr Deepjyoti Kalita, Dr Ravi Gupta, and Dr Rohit Gupta for their contribution in protocol management.

